# The World Health Organization’s Disease Outbreak News: a retrospective database

**DOI:** 10.1101/2022.03.22.22272790

**Authors:** Colin J. Carlson, Matthew R. Boyce, Margaret Dunne, Ellie Graeden, Jessica Lin, Yasser Omar Abdellatif, Max A. Palys, Munir Pavez, Alexandra L. Phelan, Rebecca Katz

## Abstract

The World Health Organization (WHO) notifies the global community about disease outbreaks through the Disease Outbreak News (DON). These online reports tell important stories about both outbreaks themselves and the high-level decision making that governs information sharing during public health emergencies. However, they have been used only minimally in global health scholarship to date. Here, we collate all 2,789 of these reports from their first use through the start of the Covid-19 pandemic (January 1996 to December 2019), and develop an annotated database of the subjective and often inconsistent information they contain. We find that these reports are dominated by a mix of persistent worldwide threats (particularly influenza and cholera) and persistent epidemics (like Ebola virus disease in Africa or MERS-CoV in the Middle East), but also document important periods in history like the anthrax bioterrorist attacks at the turn of the century, the spread of chikungunya and Zika virus to the Americas, or even recent lapses in progress towards polio elimination. We present three simple vignettes that show how researchers can use these data to answer both qualitative and quantitative questions about global outbreak dynamics and public health response. However, we also find that the retrospective value of these reports is visibly limited by inconsistent reporting (e.g., of disease names, case totals, mortality, and actions taken to curtail spread). We conclude that sharing a transparent rubric for which outbreaks are considered reportable, and adopting more standardized formats for sharing epidemiological metadata, might help make the DON more useful to researchers and policymakers.

## Introduction

The first few days are often the most important to prevent outbreaks from becoming epidemics and pandemics; by warning the world early, countries can access resource stockpiles, funding mechanisms, and decision support tools; coordinate epidemic response and surveillance across borders; and prepare the public for imminent danger. All of these have the potential to be accelerated in the internet era, but only if institutions leverage digital communications to share accurate and timely information.

The Disease Outbreak News (DON) is a public, online reporting system that serves as the only official outbreak record maintained by the World Health Organization (WHO). This system has been used since 1996, and shares event-based information provided by countries and other partners, which is organized by the WHO into prose that describes outbreak circumstances. Usually, these reports identify the geographic region affected; a known or suspected disease, or a syndromic event of unknown etiology; and some information about response, such as whether samples have been submitted for laboratory confirmation. This information helps international actors stay informed on health-related events and emergencies around the globe, and is also sometimes used by researchers as a public “database” of outbreaks (e.g., [1–3]). However, the DON is not a comprehensive record of every known disease outbreak in the world, and the internal rubric used by the WHO to decide which epidemiological data and situation updates to share is far from transparent. Over the nearly three decades that the DON has been in use, the content and subject matter of these reports may have changed in subtle ways, particularly as obligations have also changed (e.g., when the revised International Health Regulations (IHR) were adopted in 2005). Moreover, the analytic utility of the DON is severely hampered by its unstructured, prose-based system: each report is given on a separate webpage, and—while these pages are indexed by search engines like Google—there is minimal scaffolding or standardized metadata to access reports based on more structured queries. From the present interface, it is challenging to quickly access information or conduct analyses.

To support global health research that uses and interrogates the DON as both a record of outbreaks and a library of documents in its own right, we compiled basic information about reports into a standardized database. Here, we present a stable version of this resource including newly-structured metadata for each entry, collated in a machine-readable format, and show three “vignettes” that highlight how researchers can ask questions with these data. We conclude by discussing several of the limitations identified while curating this database, and suggest directions for possible improvements to the DON itself.

## Materials and methods

Our goal was to develop a metadata “skeleton” that stores key information about DON reports. To do so, we reviewed a total of 2,789 reports from the first 25 years of the DON (1996–2019), an interval that spans the start of their adoption as a formal system through the earliest days of the COVID-19 pandemic. We found that the data reported are often inconsistent within or between reports. For example, reports variously use all of the terms ‘Ebola,’ ‘Ebola Haemorrhagic Fever,’ ‘EHV,’ ‘Ebola Virus Disease,’ and ‘EVD’ to refer to the same clinical disease presentation. Similarly, geographic information may be inconsistent or unreliable: for example, DON reports often switch between spellings of cities or provinces (e.g., certain cities in Egypt appeared in consecutive reports with several different spellings).

Given the challenges of textual data sources, we adopted a mixed system of standardized and subjective descriptors in our dataset. First, we standardized as many basic variables as possible that describe the identity and subject matter of a report:

- **The identity of the report itself:** Each report is stored at its own web address (URL). In the early days, these followed one standardized format (e.g., report 1996-01-22f would be found at www.who.int/csr/don/1996_01_22f/en/ and would be the sixth report published on January 22, 1996). In later years, these have been changed to a system that instead directly represents the headline (e.g., “5-march-2019-carbapenem-resistant-p-aeruginosa-mex” for DONid DON-2019-03-05: “Carbapenem-resistant Pseudomonas aeruginosa infection - Mexico”). For each report, we provide a unique identification number (“*DONid*”) that follows the original convention (even after it was no longer adopted). Headlines are given (“*Headline*”), and usually provide unstandardized information about the year, location, and disease in question. Sometimes, these give easily interpreted names for both diseases and countries (e.g., “Plague - Madagascar”) while other times they may deviate from this template (e.g., “1997 - Viral meningitis in Gaza”, “1996 - Rwanda repatriation movement”, “1996 - Global Cholera Update 2”, “Update 73 - No new deaths, but vigilance needed for imported cases”, “Human influenza activity remained low in most countries”). If a report contains multiple countries or multiple pathogens, we split these into separate rows with the same DONid, so that metadata corresponds appropriately (see below). We also report the date a DON was published (“*ReportDate*”) and the current URL (“*Link*”). In the event that reports for certain types of outbreaks spanned two or more calendar years, outbreaks were labeled using the start year of the multi-year event.
- **The disease:** Given that the DON database is likely to be used as a database of outbreak history, we chose to implement some standardization in this field, including retrospective assignments where they improved clarity. Wherever possible, we have adopted consistent names for diseases throughout the database, and applied these names in some cases in place of temporary names that fell out of favor (e.g., DON-2012-09-23: “Novel coronavirus infection in the United Kingdom” is recorded in our dataset as MERS-CoV). We use a two-tier system to address some nuances within naming conventions: for example, all “Influenza A” values in the *DiseaseLevel1* field are accompanied, if possible, by more specific information on the HXNX subtype in the *DiseaseLevel2* field. In some cases, a report may only describe an anomaly detected by syndromic surveillance; sometimes, outbreaks are never retrospectively resolved down to a causative agent. We adopt a set of six standardized “Syndromic” categories (cardiovascular, diarrhoeal, gastrointestinal, haemorrhagic, neurological, and respiratory) for these reports, and give additional information as necessary at the second level. For example, DON-1996-10-15-a (“1996 - Outbreak of viral meningitis in Romania”) is recorded as “Syndromic: neurological” at the first level of resolution, and “Meningitis: viral” at the second.
- **The geographic area:** Most reports provide a country in the headline (“*Country*”), which we matched to 3-letter ISO country codes (“*ISO*”). We labeled travel cases based on the reported location, unless genome sequencing of the pathogen was reported as showing a clear geographic origin of the outbreak. In certain cases, if events spanned multiple countries, all countries were listed in the labeling; however, if a report labeled an outbreak as a global outbreak, or more than 10 countries were listed in the DONs reports, we labeled the event as “Global.” Additional regional locations used by the DON headlines that we preserved include “Americas,” “West Africa,” “African Meningitis Belt,” “Asia,” “Northern Hemisphere,” and “Central America.” Where possible, we also provide more detailed but unstandardized information on the reported geography (“*OutbreakEpicenter*”), based on the report text.

In addition to this basic information, we include a handful of metadata that describes additional information contained in the reports, that are necessarily—like that information—even more subjective and unstandardized. This includes:

- **Raw epidemiological data:** Where possible, we record basic descriptors on case totals and associated mortality, across five fields: *CasesTotal, CasesSuspected, CasesProbable, CasesConfirmed*, and *Deaths*. In some cases, these may report rough estimates (e.g., “>200000” total cases of dengue fever in DON-1996-01-22-b: “1996 - Dengue in the Americas”, including “>5500” cases of dengue haemorrhagic fever.) We also note whether the outbreak describes a mass gathering event as part of the epidemiological circumstances, including a handful of cases where this is contraindicated (*MassGathering*).
- **The timing of outbreak progression:** For a set of different stages in outbreak progression, we record a mix of whether a particular event has been noted to occur and/or as much information is available matching that date. That includes the declared start date of the outbreak (*OutbreakStartYear, OutbreakStartMonth, OutbreakStartDay*); whether a report is specifically describing an outbreak’s detection (*OutbreakDetection*) and when (*OutbreakDetectionYear, OutbreakDetectionMonth, OutbreakDetectionDay*); whether or not an outbreak had been verified (*OutbreakVerification*) and if so when (*OutbreakVerificationYear, OutbreakVerificationMonth, OutbreakVerificationDay*); whether there was laboratory confirmation of a suspected pathogen (*LabConfirmation*), and if so, as of when (*LabConfirmationYear, LabConfirmationMonth, LabConfirmationDay*); whether national or sub-national authorities had been notified by that point (*NotificationLocal*), and if so, the first reported date (*NotificationLocalYear, NotificationLocalMonth, NotificationLocalDay*); whether the World Health Organization had been notified or not (*NotificationWHO*), which may sometimes be indicated by requests for confirmation, and the date of notification if so (*NotificationWHOYear, NotificationWHOMonth, NotificationWHODay*); whether public communication was reported to have been undertaken by public health agencies (*PHCommunication*) and if so when (*PHCommunicationYear, PHCommunicationMonth, PHCommunicationDay*); whether public health interventions had been undertaken (*PHIntervention*), and if so when (*PHInterventionYear, PHInterventionMonth, PHInterventionDay*); whether there was any report of community resistance to intervention (*CommunityResistance*); and finally, whether a report described the outbreak as having ended (*OutbreakEnd*), and if so, when (*OutbreakEndYear, OutbreakEndMonth, OutbreakEndDay*). In many cases, the day field accompanying a reported piece of information is left empty because the report lacks specifics; more infrequently, the month field is as well.
- **Any additional information:** An unstandardized text field is reported from the researchers who coded the data (“*Notes*”) with any potentially relevant information about report content or quality control issues.

As one final observation about data quality, we note that we chose to truncate our collection at the end of 2019, to both represent an even sampling interval (the first 25 years of the dataset) and avoid any potential changes that came with the Covid-19 pandemic. Around the same time, the World Health Organization began to migrate reports to a new location on their website, with a new URL convention. In the process, several reports have become un-indexed (presumably in error); the last currently searchable record is a June 13, 2019 record (“Ebola Virus Disease - Uganda”) followed by a January 2, 2020 record (“Ebola - Democratic Republic of the Congo”). At present, our database may be the only record of reports that span the second half of the year—the final two reports of the year are both from December 26, 2019—and their URLs have been updated to follow the same convention, even though these links are presently dead, with the expectation these reports will return soon online.

## Results

While the total number of entries in the DON vary significantly from year to year, there has been no consistent increase or decrease over time. The number of reports published annually ranged from 59 reports in 2011 to 205 reports in 2014 (Figure 1), with an average of 116.2 reports per year. These reports are distributed very unevenly across countries and pathogens, with a substantial covariance between the two (Table 1), and the most reports occur in years with notable outbreaks (e.g., the 2014-16 epidemic of Ebola virus in West Africa). Over time, a smaller number of diseases account for a greater proportion of these reports—an unusual trend as the rate of disease emergence has grown globally in the last few decades [4–6], but one that reflects the growing duration and complexity of some epidemics. Some diseases are only reported during major events: for example, anthrax is a persistent and global disease of livestock keepers, with frequent major epidemics, but almost only occurs in the DON in the context of the 2001 bioterrorism incidents [7,8]. Similarly, West Nile virus is mostly reported during its spread through North America in the early 2000s. Other similar diseases, like plague or Rift Valley ever, are reported evenly throughout the 25-year interval. Though this tends to correspond to human-to-human transmissibility, and so epidemic concerns, that reasoning is sometimes more unclear. For example, dengue is reported evenly throughout the interval, while malaria is almost entirely reported in 1998 during two separate incidents (an epidemic in east Africa in May, and a November-December epidemic in central America following Hurricane Mitch).

**Table 1.** The top reported countries, diseases, and country-disease pairs by total number of DON reports between 1996 and 2019 (number given in parentheses).

| Top ten countries | Top ten diseases | Top ten country-disease pairs |
| --- | --- | --- |
| 1. China (262) | 1. Influenza A (776) | 1. China - Influenza A (218) |
| 2. Saudi Arabia (189) | 2. MERS-CoV (316) | 2. Saudi Arabia - MERS-CoV (179) |
| 3. Dem. Rep. of the Congo (171) | 3. Ebola virus (308) | 3. Indonesia - Influenza A (123) |
| 4. Indonesia (146) | 4. Cholera (278) | 4. Egypt - Influenza A (112) |
| 5. Egypt (115) | 5. Yellow fever (162) | 5. Dem. Rep. of the Congo - Ebola virus (105) |
| 6. Uganda (83) | 6. Meningococcal disease (123) | 6. Vietnam - Influenza A (77) |
| 7. Vietnam (81) | 7. SARS-CoV (118) | 7. Uganda - Ebola virus (57) |
| 8. Liberia (74) | 8. Polio (105) | 8. Guinea - Ebola virus (53) |
| 9. Nigeria (69) | 9. Dengue fever (58) | 9. Liberia - Ebola virus (49) |
| 10. Guinea (68) | 10. Marburg fever (52) | 10. Gabon - Ebola virus (44) |

**Figure 1.**
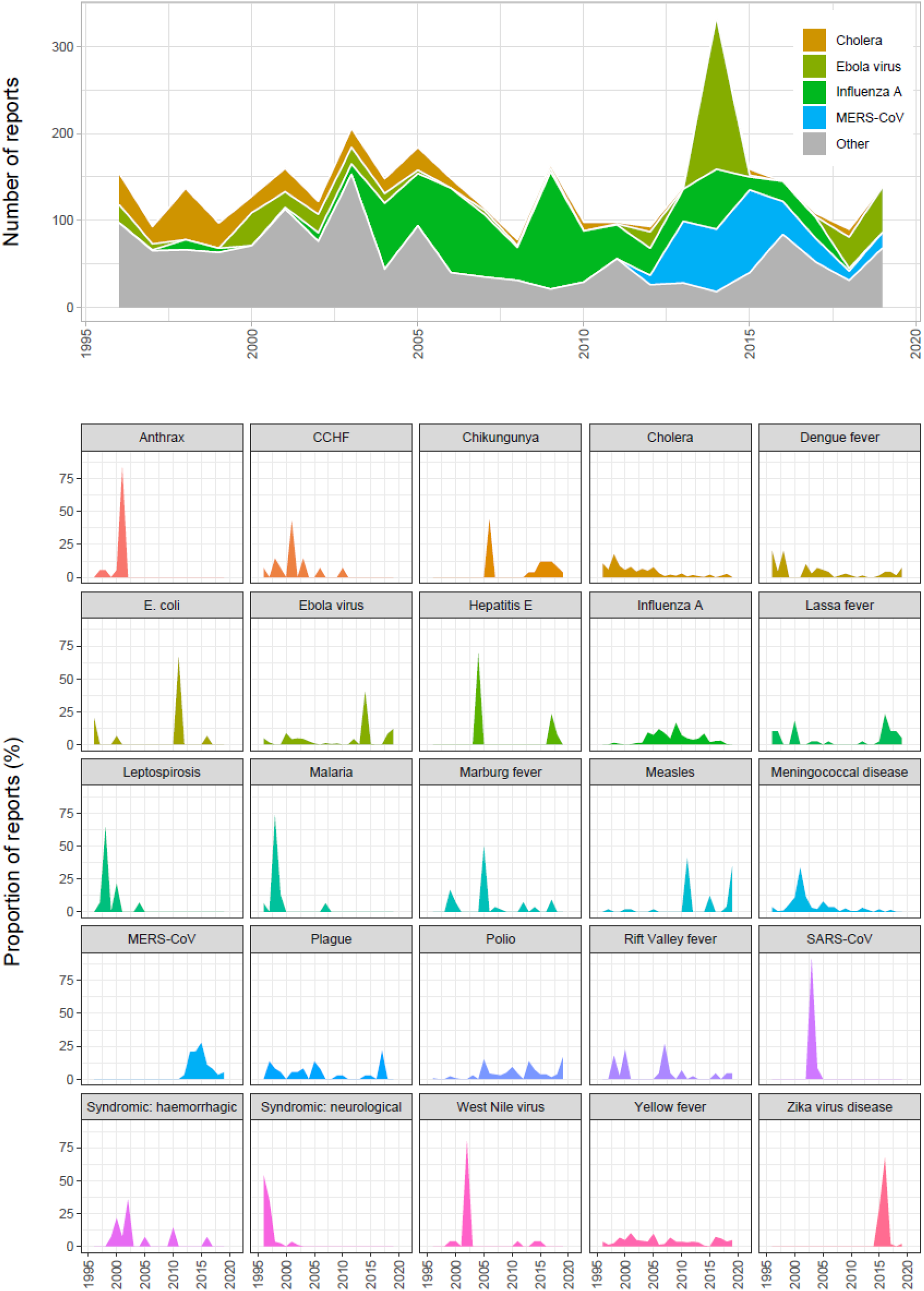
The distribution of outbreaks through time. (top) Number of reports per year, broken down to highlight the top four diseases, which are reported substantially more often than all others (see Table 1). (bottom) The proportion of total reports of a disease, across years, that occur in a given year, for the top 25 reported diseases in alphabetical order.

**Figure 2.**
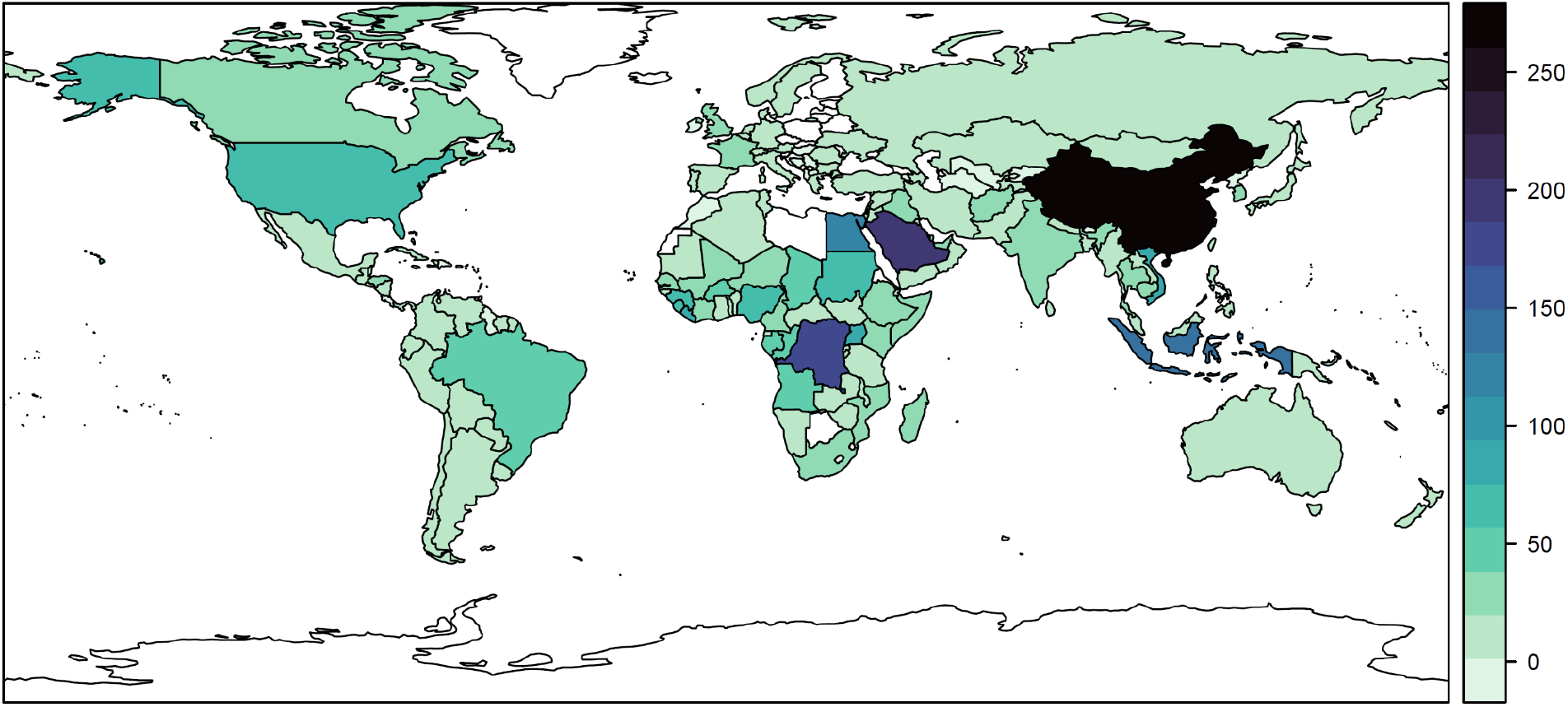
The number of reports by country (blank countries indicate no reports ever published; however, some reports are also global in scale, or describe entire regions like the Americas).

**Figure 3.**
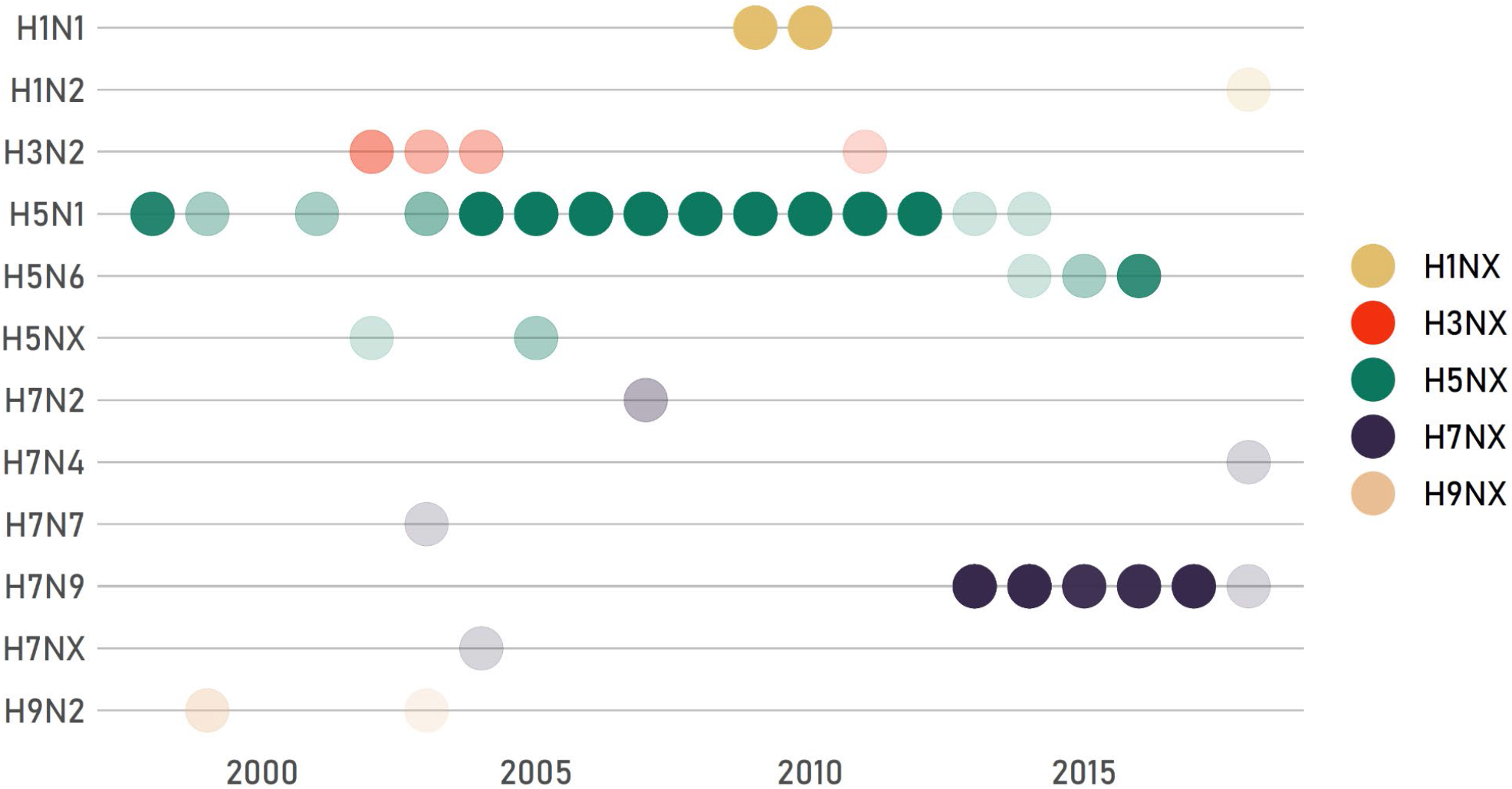
Influenza subtypes represented in the DONs shift over time. Each dot represents a year with at least one notification; opacity is proportional to the number of DON reports for that subtype in each year. Note that some reports do not specify a relevant subtype.

**Figure 4.**
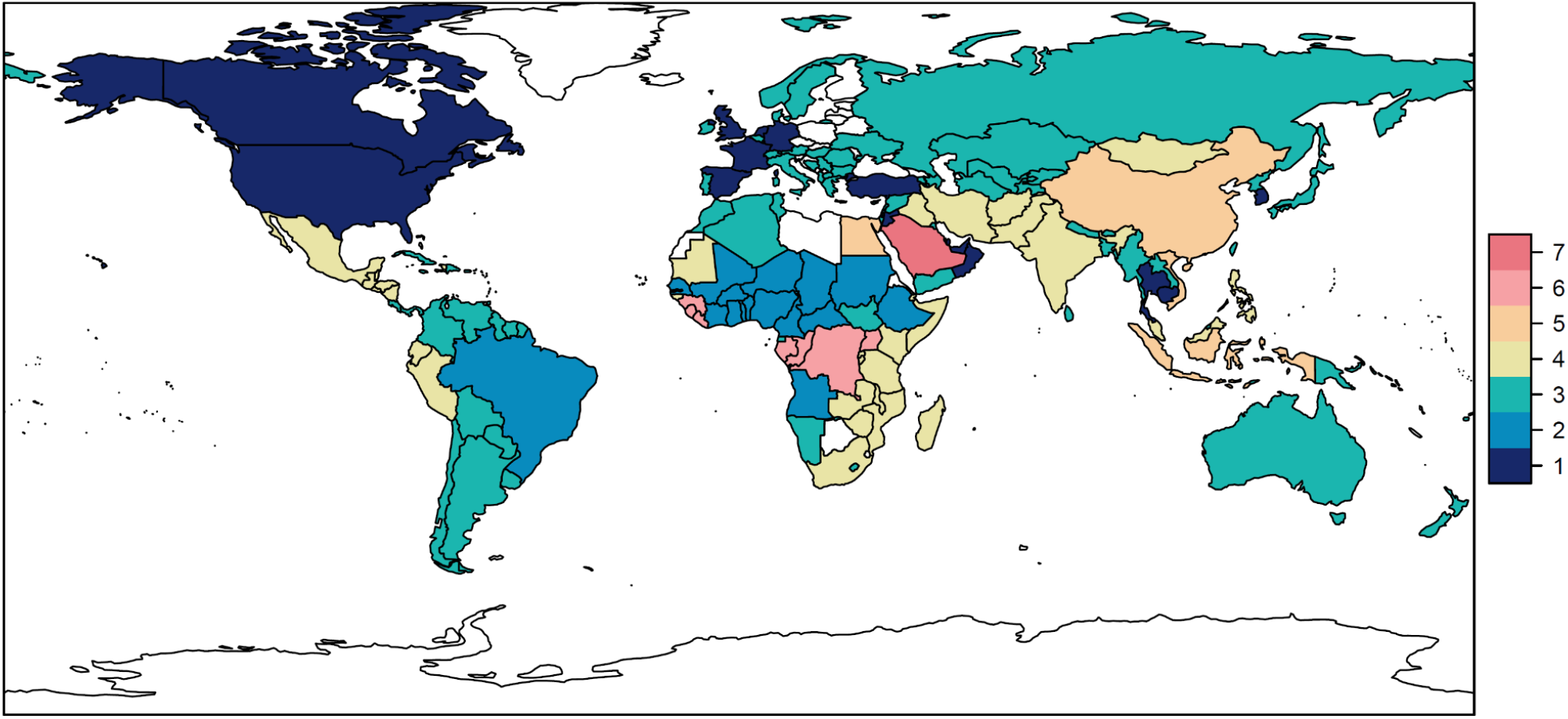
Which countries report more similar kinds of outbreaks? Seven clusters are mapped (each a different color) based on an unsupervised clustering algorithm (k-means).

The number of reports varies substantially among countries, and is strongly reflective of reporting of key pathogens like influenza and Ebola. For example, Egypt is the fifth most common country across reports, while neighboring Libya has never been associated with a single report, reflective of the collapse of its (once notoriously successful) health system after two civil wars [9,10]. Other countries that dominate reporting are China (due to influenza and, to a lesser degree, SARS-CoV reports), Indonesia (again for flu), the Democratic Republic of the Congo (for Ebola), and Saudi Arabia (for MERS-CoV).

As a final note, we found that recently, the DON has been used for fewer reports of actual “news” of global concern that do not describe an outbreak *per se*. Though this means the DON is now a more faithful record of global outbreak surveillance, it loses some of the editorial aspects that have made these reports unique insights into the World Health Organization and its inner workings. Take DON-2003-06-26: “Update 89 - What happens if SARS returns?”, which identifies five reasons that “should SARS return, it will not do so with a vengeance” (Box 1). The reasons given, in order, are: re-emergence would be taken seriously by the global community; countries can implement basic transmission-interrupting measures, as they did during the SARS outbreak; research into coronaviruses, and particularly the development of rapid tests, would improve biomedical surveillance and countermeasures; countries would choose to strengthen the World Health Organization in the months and years following the SARS outbreak; and countries both could not and would not fail to disclose another emergence event quickly, in part because “SARS is simply too big a disease to hide for long.” In retrospect, these reasons nearly perfectly correspond to the failures that were most consequential in the early COVID-19 pandemic [11]. Though these digressions within the DON complicate its use for research, their editorial slant also preserves the “thought process” of the WHO at key moments in its institutional history—opening a window into the inner workings of an institution that has, at times, struggled with transparency—and surely have historical value as such.

### How to use the DON database

To illustrate the value of these data, we show three simple analyses that researchers could undertake using these data, with different levels of effort.

#### Example 1. “Lost” outbreaks of hemorrhagic fevers

Spillover events are often undetected, even for syndromically-conspicuous diseases. Previous work has suggested that at least half of all Ebola virus spillovers are either undetected or unreported [12]. These rates could be even higher for diseases with less targeted surveillance, especially in parts of Africa where pathogen diversity is high in animal reservoirs [13], febrile illness is common and complicates syndromic detection [14], and diagnostic capacity is limited, especially for metagenomic screening to identify novel pathogens. It may be possible to identify “missing outbreaks” through retrospective analyses, and the DON could be used as a starting point for such an analysis. A total of 14 reports are recorded in our dataset as “Syndromic: haemorrhagic,” spanning 1999 to 2016, and almost entirely in African nations where haemorrhagic fevers like Ebola and Marburg pose a persistent problem (Democratic Republic of the Congo, Republic of the Congo, Gabon, and South Sudan). However, the majority of these reports offer a counter-explanation (e.g., “2000 - Acute haemorrhagic fever syndrome in Afghanistan” indicates that symptoms and circumstances were compatible with Crimean-Congo haemorrhagic fever [CCHF]; “Acute haemorrhagic fever syndrome in Timor-Leste” with dengue haemorrhagic fever; and so on). One particularly notable report, though, describes the 2015-16 outbreak of haemorrhagic fever in South Sudan that remains poorly understood. The report notes positive tests for O’nyong-nyong virus, chikungunya virus, and dengue virus, but negative tests for seven other diseases, including Ebola, Marburg, and CCHF. A retrospective study in 2019 found 11% of tested samples from the same outbreak came up positive for CCHF [15]; the DON reports could be useful to motivate similar retrospective studies. However, this utility may be diminished by how rarely the DON is used to report routine syndromic surveillance: of the 14 reports, only eight total events are described (six are updates), and the global number of unexplained outbreaks of severe febrile illness since 1996 is surely higher.

#### Example 2. Shifting significance of influenza subtypes

The global influenza surveillance system is an unparalleled network of information sharing. Countries are required to report human cases of novel influenza subtypes to the World Health Organization as part of its obligations under the International Health Regulations, and routinely submit an even greater range of data voluntarily, which are often reflected in the DON. By tracking which subtypes are described in the DON over time, researchers can track persistent spillovers of H5N1 avian influenza into human populations, a global concern that spans multiple decades, as well as the 2009 H1N1 pandemic and the more recent emergence of the H7N9 subtype in China in 2013. However, these data also show the major gaps in the DON compared to the underlying “reality” of outbreaks. For example, H1N1 outbreaks are only reported in the 2009-2010 period, but several major epidemics have occurred since, including a 2015 epidemic in India with over 30,000 cases, and a 2019 epidemic in Iran with over 4,000 hospitalizations. These additional outbreaks are unrecorded in the DON, for which influenza reports have over time become almost entirely restricted to China: one 2015 report of H7N9 in Canada, a 2018 report of H1N2 in the Netherlands, and a 2020 report of H5N1 in Laos are the only reports on influenza A outside China after 2014. The reporting criteria underlying these subjective decisions in curation are unknown.

#### Example 3. The biogeography of disease outbreaks

As a final illustration of how these data could be used for more quantitative research, we conducted a simple analysis to explore which countries report more similar kinds of diseases. To do so, we counted the number of times a given disease was reported in each country, root-transformed this matrix (given how many updates have been filed in a small number of epidemics), and sent this matrix through a k-means clustering algorithm (with an arbitrarily chosen *k* = 7 clusters). Mapping these clusters shows particular areas of global focus that are represented in the DON: one cluster (orange) is clearly driven by influenza surveillance, while another (red in Saudi Arabia) is driven by the MERS-CoV epidemic, and another (light pink) is linked to Ebola virus outbreaks. The United States, Canada, and a handful of countries in Western Europe also cluster together (dark blue)—highlighting how separate the Global North is from the rest of the world in terms of both disease burden and surveillance priorities.

## Discussion

Here, we have developed and documented a database of the WHO Disease Outbreak News, the only official record of outbreaks maintained by the organization. By indexing key information about these reports and making the semi-structured data publicly available, we hope to support future research that uses them both to investigate outbreak dynamics over time, and where possible, the meta-textual elements of when, how, and why outbreaks are reported. In future versions of the dataset, we hope to add additional information that might continue to improve this resource. In particular, mapping the DON onto an independent record of outbreak events and identifying which reports are linked—a property that is often, but not always, possible to reconstruct from the report websites—would make the dataset a clearer record of outbreaks, rather than simply reporting.

When the DON were adopted in January 1996, they marked a significant shift in the WHO’s move from facsimile to electronic outbreak and rumor information sharing [16]. This empowered global outbreak surveillance, while developing an archive for subsequent research. As highlighted in the methods, during the Covid-19 pandemic, WHO’s website underwent substantial updates. In this process, approximately six months of DON reports were not relocated to the new site, and are not available at the original links. However, we managed to capture this data in our dataset before it became lost. From a governance perspective, it is critical that disease outbreak information, like the DON, are preserved through technological advances – from telegram to facsimile to an email listserv to a publicly accessible website.

The process of curating the DON into a workable database has also raised significant questions about its utility as its stated purpose: a formal system for documenting the world’s outbreaks. There is no standard format or structure for DON reports; they vary greatly in length, and the information included has changed over time, and is often contingent on the specific disease, country, level of emergency, and report author. While the lack of structure may improve flexibility, and almost certainly reflects the information reported to the WHO, these inconsistencies make it difficult to quickly identify important information. For example, many reports contain useful basic information about case totals or key dates, but those data are rarely standardized or reusable for quantitative analysis; the inclusion of data tables, charts or links in reports solves this problem partially, but their inclusion is infrequent, and does not adhere to specific patterns or methodologies. The reliability of the data included is also inconsistent and sometimes troubling. For example, some reports are not published until months after initial reporting of the outbreak to relevant authorities (DON-2011-07-22: “Cholera outbreaks in the Democratic Republic of Congo (DRC) and the Republic of Congo”, which describes outbreaks in these two countries reported in March and June 2011 respectively). Similarly, a report on avian influenza in Vietnam published on January 11, 2009 reports (presumably erroneously) that an individual first developed symptoms on January 28, 2009 and was hospitalized on January 31, 2009, several weeks after the report was published (DON-2009-01-11: “Avian influenza - situation in VietNam - update”). Other reports contain similar errors and chronological inconsistencies, such as the same report published twice on consecutive days (DON-2009-05-26: “Influenza A(H1N1) - update 39” and DON-2009-05-27-a: “Influenza A(H1N1) - update 39”).

All of these aspects complicate the use of the DON (and our database of the reports) as a textual resource describing the global history of infectious disease outbreaks. It should therefore be unsurprising that researchers have often turned to other informal or privatized sources of information. For example, the International Society for Infectious Diseases’ Program for Monitoring Emerging Diseases (ProMED-mail; https://promedmail.org/) and HealthMap (https://www.healthmap.org/en/) are freely-available data sources, and the GIDEON database (https://www.gideononline.com/) provides access to global infectious disease surveillance data for a subscription fee. All of these sources are used for research and action— ProMed in particular was widely hailed as one of the earliest sources of public information on the Covid-19 pandemic (or, more precisely, the pneumonia outbreak in Wuhan)—but are unofficial and for the purposes of outbreak reporting, fall under Article 9 of the IHR, which permits WHO to receive “other reports” and to seek verification from countries under Article 10, rather than official notifications from States Parties under Article 6 of the IHR.

If the purpose of the DON is to share the information submitted by Member States, it would be valuable to improve the transparency of the process. Given that the reported outbreaks are only a subset of those that occur in ProMed or similar sources—and a small subset of those that qualify as notifiable under the IHR— it would be valuable for the WHO to publish the internal criteria outlining what events merit a report (or, if those criteria are not formalized, to develop them). Moreover, at present, the WHO does not publish any secondary information about the reports, such as the authors of reports or the criteria for publication. Including information detailing how reports are prepared and compiled would further improve transparency, not just for the World Health Organization but for efforts to understand States’ compliance with international governance [11].

If the DON are indeed intended to be the global database of record for infectious disease outbreaks, further changes would significantly improve their usability. Specifically, we believe that the WHO could improve DON reports by implementing a consistent and systematized reporting format related to epidemiological data. An appropriate place to start could be the criteria put forward by Smolinski and colleagues [17]: case total tables organized by defined probable, suspected and confirmed cases; and subsections for appropriate or priority contextual factors, such as meteorology or climate hazards, community resistance, conflict, migration, or mass gatherings. Doing so would make information easier to find, ensure consistent reporting, and could allow for the reports to become machine-readable, and thus a more accessible source of information. This would also facilitate States Parties’ compliance with obligations to share public health information under the IHR when making an Article 6 notification. Establishing these parameters and guidelines would improve the transparency and standardization of the reporting process, could act to improve confidence in the reports themselves as the single authoritative collection of disease outbreaks, and might allow for it to be more easily adapted and used for analytic purposes. In the absence of these changes, more standardized sources like the Global.health (https://global.health/) project’s epidemiological line data may outpace the WHO’s own capacity to support outbreak research in global health.

## Data Availability

The DON database is available on Github, with all scripts required to reproduce the study, at https://github.com/cghss/dons. In addition to this manuscript, the repository itself can be cited directly as Zenodo DOI: 10.5281/zenodo.6374495.

## Acknowledgements

This work was funded by the Open Philanthropy Project. CJC and ALP were additionally supported by the Carnegie Corporation of New York (Grant G-21-58414). The funders had no role in the study design; data collection, analysis, and interpretation; preparation of the manuscript; decision to publish. We also thank Emily Woodrow and Harshini Tammareddy for additional support on data development.

## Author Contributions

RK conceived of the project. All authors contributed to the design of the project, the collection of data, and the writing of the manuscript. CJC contributed analysis and visualization.

## Display Items

Box 1. The below text is a directly quoted passage from “Update 89 – What happens if SARS returns?” (see discussion in the main text).

*…WHO has good reason to believe that, should SARS resurface later this year, the global impact will be milder than experienced during the initial global emergency. Five reasons support this view*.

*First, the world’s public health systems have demonstrated their capacity to move quickly into a phase of high alert. The prompt detection and isolation of imported cases in African and India are good examples of both the level of vigilance and its effectiveness in preventing further spread. Some of the former SARS hotspots, including Hong Kong and Singapore, plan to maintain a high level of vigilance, supported by measures for screening and detection, until at least the end of the year*.

*Second, the world knows what to do. Control measures, though centuries old, have demonstrated their capacity to completely halt outbreaks, as most recently seen in Singapore, Hong Kong, and Beijing*.

*Third, the intensive research effort currently under way can be expected to improve scientific understanding of SARS and yield better control tools, most notably a rapid and reliable point-of-care diagnostic test*.

*Fourth, resolutions adopted during the May World Health Assembly have strengthened WHO’s capacity to respond to outbreaks in important ways. In effect, these resolutions allow WHO to move from a passive reliance on official government notifications to a proactive role in warning the world as soon as evidence indicates that an outbreak poses a threat to international public health*.

*Finally and perhaps most importantly, SARS has underscored the importance of immediately and fully disclosing cases of any disease with the potential for international spread. In the present climate of opinion, influenced by the lessons learned from SARS, it appears unlikely that any country would choose to conceal cases, should SARS resurface. In addition, SARS is simply too big a disease to hide for long*.

*For these reasons, WHO is optimistic that, should SARS return, it will not do so with a vengeance*.

## References

1. Lugo-Robles R, Garges EC, Olsen CH, Brett-Major DM. Identifying Nontraditional Epidemic Disease Risk Factors Associated with Major Health Events from World Health Organization and World Bank Open Data. Am J Trop Med Hyg. 2021;105: 896–902.

2. Warsame A, Murray J, Gimma A, Checchi F. The practice of evaluating epidemic response in humanitarian and low-income settings: a systematic review. BMC Med. 2020;18: 315.

3. Oppenheim B, Gallivan M, Madhav NK, Brown N, Serhiyenko V, Wolfe ND, et al. Assessing global preparedness for the next pandemic: development and application of an Epidemic Preparedness Index. BMJ Glob Health. 2019;4: e001157.

4. Jones KE, Patel NG, Levy MA, Storeygard A, Balk D, Gittleman JL, et al. Global trends in emerging infectious diseases. Nature. 2008;451: 990–993.

5. Smith KF, Goldberg M, Rosenthal S, Carlson L, Chen J, Chen C, et al. Global rise in human infectious disease outbreaks. Journal of The Royal Society Interface. 2014. p. 20140950. doi:10.1098/rsif.2014.0950

6. Woolhouse M, Scott F, Hudson Z, Howey R, Chase-Topping M. Human viruses: discovery and emergence. Philos Trans R Soc Lond B Biol Sci. 2012;367: 2864–2871.

7. Carlson CJ, Kracalik IT, Ross N, Alexander KA, Hugh-Jones ME, Fegan M, et al. The global distribution of Bacillus anthracis and associated anthrax risk to humans, livestock and wildlife. Nat Microbiol. 2019;4: 1337–1343.

8. Carlson CJ, Getz WM, Kausrud KL, Cizauskas CA, Blackburn JK, Bustos Carrillo FA, et al. Spores and soil from six sides: interdisciplinarity and the environmental biology of anthrax (Bacillus anthracis). Biol Rev Camb Philos Soc. 2018;93: 1813–1831.

9. Sullivan R, McQuinn B, Purushotham A. How are we going to rebuild public health in Libya? J R Soc Med. 2011;104: 490–492.

10. Iwendi GC, Alsadig AM, Isa MA, Oladunni AA, Musa MB, Ahmadi A, et al. COVID-19 in a shattered health system: Case of Libya. Journal of Global Health. 2021. doi:10.7189/jogh.11.03058

11. Singh S, McNab C, Olson RM, Bristol N, Nolan C, Bergstrøm E, et al. How an outbreak became a pandemic: a chronological analysis of crucial junctures and international obligations in the early months of the COVID-19 pandemic. The Lancet. 2021. pp. 2109–2124. doi:10.1016/s0140-6736(21)01897-3

12. Glennon EE, Jephcott FL, Restif O, Wood JLN. Estimating undetected Ebola spillovers. PLoS Negl Trop Dis. 2019;13: e0007428.

13. Olival KJ, Hosseini PR, Zambrana-Torrelio C, Ross N, Bogich TL, Daszak P. Erratum: Host and viral traits predict zoonotic spillover from mammals. Nature. 2017;548: 612.

14. Glennon EE, Jephcott FL, Oti A, Carlson CJ, Bustos Carillo FA, Hranac CR, et al. Syndromic detectability of haemorrhagic fever outbreaks. bioRxiv. medRxiv; 2020. doi:10.1101/2020.03.28.20019463

15. Bower H, El Karsany M, Alzain M, Gannon B, Mohamed R, Mahmoud I, et al. Detection of Crimean-Congo Haemorrhagic Fever cases in a severe undifferentiated febrile illness outbreak in the Federal Republic of Sudan: A retrospective epidemiological and diagnostic cohort study. PLoS Negl Trop Dis. 2019;13: e0007571.

16. Dzenowagis J. Using electronic links for monitoring diseases. World Health. 1997;50: 8–9.

17. Smolinski MS, Crawley AW, Olsen JM. Finding Outbreaks Faster. Health Secur. 2017;15: 215–220.

